# Gene-Burden Meta-Analysis of 748,879 Individuals Identifies LGI1-ADAM23 Protein Complex Association with Epilepsy

**DOI:** 10.1101/2024.12.30.24319794

**Authors:** Jessica Castrillon Lal, Costin Leu, Christian M. Boßelmann, Alina Ivaniuk, Eduardo Pérez-Palma, Dennis Lal

## Abstract

Epilepsy affects over 50 million individuals globally and has a substantial genetic component that remains to be completely understood. Traditional studies have focused on severe, early-onset cases enrolled through clinical or research settings. Recent biobank-based approaches, leveraging large-scale population datasets, offer opportunities to explore genetic associations in broader epilepsy phenotypes, including milder, later-onset forms. We analyzed data from over 750,000 individuals across the UK Biobank, All of Us, and Massachusetts General Brigham Biobank, including 20,026 individuals with epilepsy. Rare coding variant burden testing revealed a significant association with *LGI1*, a known epilepsy gene. Other top-ranked genes, including *GABRG2, ATP1A3*, and *ADAM23*, demonstrated strong enrichment for epilepsy-associated variants. Notably, *ADAM23*, previously linked to epilepsy in dogs and strong brain expression in humans, emerged as a novel candidate, potentially contributing to human epilepsy through its direct interaction with *LGI1*. Phenome-wide analyses highlighted the pleiotropic effects of epilepsy genes, with *LGI1* and *ADAM23* predominantly associated with epilepsy, while other genes such as *KRIT1, TSC1*, and *TSC2* exhibited broader systemic involvement. Our study shows the potential of population-scale genomic data and suggests that integrating these datasets with deep phenotyping will uncover more novel insights into epilepsy genetics in the future.

## Introduction

Epilepsy is a common neurological disorder that affects over 50 million individuals worldwide and is characterized by a significant but incompletely understood genetic component^1-3^. Conventional clinical research strategies, e.g., gene burden association studies on clinically ascertained cohorts, have predominantly focused on severe, early-onset forms, e.g., developmental and epileptic encephalopathies (DEEs), elucidating key pathways such as ion channel dysfunction, synaptic communication, and mTOR signaling ^2, 4, 5^. In parallel, large-scale research efforts, such as the Epi25 Collaborative, have demonstrated the value of deeply phenotyped datasets in uncovering subtype-specific genetic associations, including ultra-rare variant contributions across DEEs, genetic generalized epilepsies (GGE), and non-acquired focal epilepsies (NAFE)^2^. These recent large-scale research cohort studies underscore the critical role of integrating genetic data with detailed phenotypic characterization to reveal both shared and distinct molecular mechanisms across epilepsy subtypes^2^. A potential major barrier to discovering new genes in epilepsy is that many potential genetic patients are excluded because they do not meet the commonly used clinical criteria—such as those defined by the International League Against Epilepsy syndrome definitions (https://epi-25.org/epi25-eligibility-criteria).

Biobank-based approaches, which draw on population-scale datasets, offer new opportunities to identify genes linked to more broadly defined epilepsies, as most phenotype classifications are deliberately broadly based on billing codes rather than strict clinical criteria. The large sample size increases statistical power to detect associations with mild or low-penetrance genetic variants that might not emerge in smaller studies, and are considered to have less of a Mendelian genetic contribution^6-8^. Meanwhile, population-based studies, like those leveraging the UK Biobank and other global resources, have revealed the power of pan-ancestry analyses to uncover rare genetic associations across diverse populations^9^. The massive cohort sizes of recent cross-biobank meta-analyzes have shown great success in identifying novel disease-associated genes and may also enable the identification of novel genes associated with epilepsy^9^.

Here, we explore publicly available genomic resources for exome-wide gene–epilepsy associations in a cohort of more than 750,000 individuals, including 20,026 people with epilepsy, drawn from three biobank resources: All of Us (AoU), the UK Biobank (UKB), and the Massachusetts General Brigham Biobank (MGB). By leveraging the scale and diversity of these datasets, we can identify both well-established and previously unknown epilepsy-associated genes.

## Methods

### Dataset, origin and composition

The dataset utilized in this analysis originates from a comprehensive pan-ancestry rare coding variant study across three major biobanks: the UK Biobank (UKB), the All of Us (AoU) Research Program, and the Mass General Brigham (MGB) Biobank^9^ that utilized either exome or genome sequencing to identify rare variants. We accessed publicly available gene-level association results through a website provided by the authors in Dec 2024 (https://hugeamp.org:8000/research.html?pageid=600_traits_app_home). A detailed description of the cohort and methods can be found in the referenced study^9^. Our work focuses on re-analyzing and interpreting these results in the context of epilepsy in combination with additional analyses, leveraging our domain-specific expertise. In brief, the combined dataset includes sequencing data for 748,879 individuals: 454,162 from UKB (whole-exome sequencing), 242,902 from AoU (whole-genome sequencing), and 51,815 from MGB (whole-exome sequencing). Participants were grouped into principal ancestry categories using genetic similarity to 1000 Genomes reference samples, with 20.7% of individuals classified as non-European ancestry (notably African, Admixed-American, East Asian, and South Asian). The cohort is expected to primarily include adults with a spectrum of epilepsy types, ranging from mild to moderate cases, with a likely underrepresentation of severe epilepsy syndromes and individuals with significant cognitive disabilities. This reflects the recruitment biases of biobanks and hospital datasets, which tend to include more generalizable populations and fewer individuals with profound neurological impairments.

### Phenotypic Endpoints and association testing

Phenotypic endpoints were derived from International Classification of Disease (ICD) codes mapped into 1,866 phecodes, and subsequently pruned to 601 disease phenotypes using hierarchical clustering to minimize redundancy. Disease prevalence was standardized across cohorts, with adjustments for potential differences in coding systems (ICD in UKB vs. ICD-CM in AoU and MGB). The only epilepsy-related phecode that was mappable across all three studies was #345 ‘Epilepsy, recurrent seizures, convulsions’, annotated for 20,026 people. The final dataset underwent gene-based burden testing for rare variants (minor allele frequency <0.1%) across predefined masks (loss-of-function variants and missense variants with functional scores), followed by meta-analysis with corrections for sample overlap. To assess pleiotropic effects, we utilized PheWAS results available through the online resource. These analyses examined phenotypic associations for selected epilepsy-related genes across 601 disease endpoints. All statistical analyses were conducted by the original study authors. Our contribution was limited to summarizing and interpreting the associations for epilepsy and related traits.

### Evaluation of Gene Burden Association Tests

To evaluate the performance of our gene burden association tests, we conducted a statistical analysis to assess the probability of observing established epilepsy genes among the top-ranked results. Of the approximately 20,000 human protein-coding genes, 1,016 are known to be associated with epilepsy to date based on the Genes4Epilepsy database (https://github.com/bahlolab/genes4epilepsy, accessed December 2024)^10^. Using a binomial test, we compared the observed number of established epilepsy genes (4 out of the top 5 associations) to the expected probability under random chance, calculated as the proportion of epilepsy-associated genes (1,017/20,000). A one-sided t-test was performed to determine whether the observed enrichment was statistically significant.

### Gene Expression Analysis

Gene expression data for *ADAM23* was analyzed to examine tissue-specific and developmental expression patterns across humans and other species. In humans, tissue-specific expression was quantified using transcript per million (TPM) values obtained from the GTEx database (https://www.gtexportal.org/home/gene/ADAM23). Developmental expression profiles in humans (ENSG00000114948), mice (ENSMUSG00000025964), rats (ENSRNOG00000012424), and chickens (ENSGALG00000008582) were retrieved from the Evo-Devo App (https://apps.kaessmannlab.org/evodevoapp/). Reads per kilobase of transcript per million mapped reads (RPKM) values were used to measure expression levels across pre- and postnatal developmental stages.

### Other Bioinformatic Resources Utilized

Rare variant constraint information for genes, including *ADAM23*, was retrieved from the gnomAD V4 database (https://gnomad.broadinstitute.org/gene/ENSG00000114948?dataset=gnomad_r4). Enrichment analyses for pathway overrepresentation, were conducted using the KEGG pathway annotation via the Enrichr platform (https://maayanlab.cloud/Enrichr/).

## Results

In this meta-analysis of 748,879 individuals, the phecode #345 “Epilepsy, recurrent seizures, convulsions” was annotated for 20,026 individuals (Figure 1). The strongest rare variant burden association, and the only one surpassing the multiple testing correction threshold, was observed for *LGI1* (*P* = 1.85 x 10^-7^) under the loss-of-function plus rare missense variant model (Figure 1). *LGI1* is a well-established gene associated with autosomal dominant lateral temporal lobe epilepsy. Given that only 1,016 of ∼20,000 human genes have been associated with epilepsy to date (Genes4Epilepsy, see Methods), the probability of observing this level of enrichment by random chance was (*P* = 3.2 x 10^−5^). While no other genes reached statistical significance after multiple testing correction, the cross-biobank results demonstrated strong validity and appeared well-calibrated, as four additional established epilepsy-associated genes ranked among the top five associations (Figure 1). These four additional established epilepsy-associated genes among the top five included *GABRG2* (*P* = 3.99 x 10^-6^) and *ATP1A3* (*P* = 1.44 x 10^-5^), both under the loss-of-function plus rare missense variant model. Interestingly, the fourth strongest association was found for truncating variants in *ADAM23* (*P* = 5.54 x 10^-6^, Loss of function model) a gene constrained for truncating variants in humans (gnomAD, see methods) that has previously been associated with epilepsy in dogs and is a direct interaction partner of *LGI1*^*11*, *12*^. Expression analysis further demonstrated that *ADAM23* is highly enriched in the brain across multiple species, with a sharp increase in expression postnatally (Figure 2). This developmental regulation aligns with its known role in epilepsy pathophysiology ref?. Additionally, ADAM23 expression in humans was observed to be strongly brain-specific, with negligible expression in other tissues (Figure 2A). Additional well-established genes in the top 50 include *KRIT1, TSC1* and *TSC2* and *DEPDC5* and *GRIN2A* (Figure 1). This group of genes indicates a strong overrepresentation of the mTOR signaling pathway (*P*_adjusted_= 9.26 x 10^-4^, KEGG pathway hsa04150, see Methods).

**FIGURE 1.**
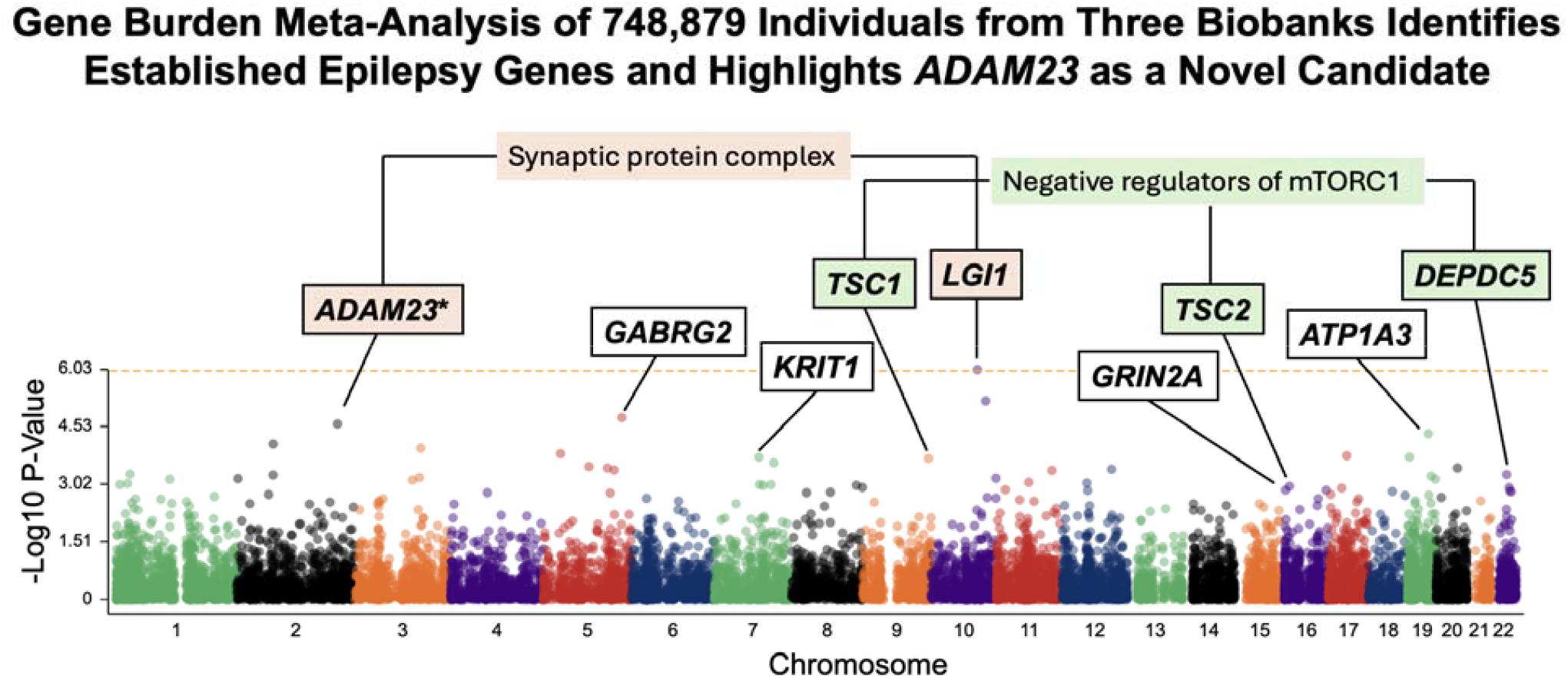
Association Results for Gene Burden Test, Manhattan plot depicting the results of the gene burden association test. The y-axis represents the -log10(p-value) of the associations, indicating the statistical significance of each gene’s burden test. The x-axis is organized by chromosomal positions, with each data point corresponding to a specific gene. Genes identified as well-established human epilepsy genes and ranking among the top associations are labeled. Notably, ADAM23—an established canine epilepsy gene and a direct protein-protein interaction partner of LGI1, the gene with the strongest association in this study—is highlighted with an asterisk (*).

**FIGURE 2.**
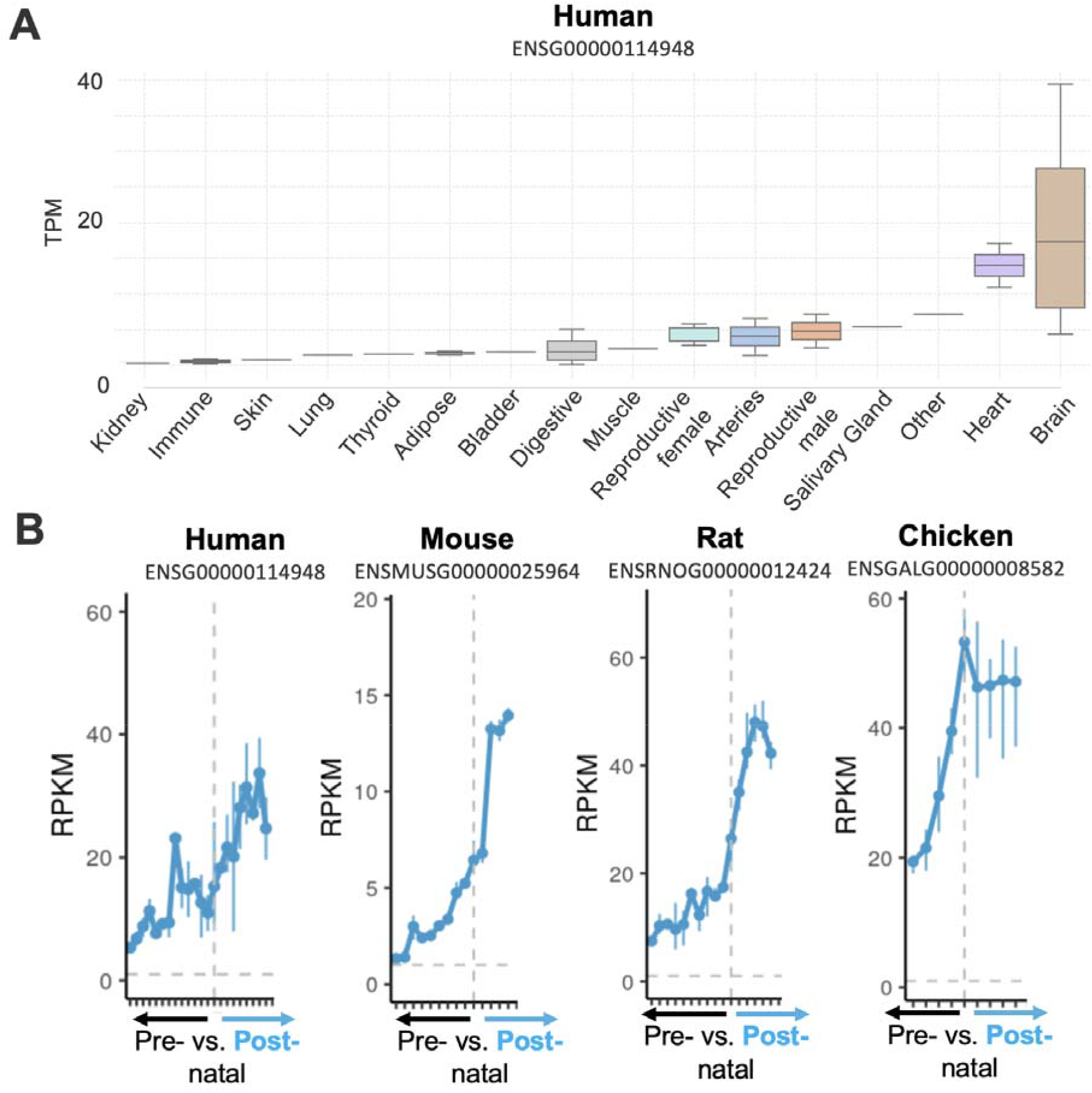
ADAM23 is mainly postnatal brain expressed in humans. A) ADAM23 human gene expression across organs of adult donors. B) ADAM23 gene developmental gene expression in human, mouse, rat and chicken. Data visualization included boxplots to depict human tissue-specific expression and line plots to illustrate developmental trends across species.

To assess the specificity of the observed associations, we performed PheWAS analyses for gene-burden signals across >600 phecodes (Table S1). *LGI1, ADAM23, GABRG2*, and *ATP1A3* showed epilepsy/recurrent seizures as the strongest association, consistent with their established roles in epilepsy. For *KRIT1*, the strongest associations were observed for hemangioma and lymphangioma, surpassing epilepsy/recurrent seizures in significance. For *TSC1* and *TSC2*, the strongest associations were observed for other and unspecified congenital anomalies, with atrial fibrillation/flutter also reaching significance for *TSC2*. In both cases, epilepsy/recurrent seizures remained significant but secondary. *DEPDC5* showed stronger associations with nystagmus and irregular eye movements, Herpes zoster, and malignant uterine neoplasms, with epilepsy/recurrent seizures ranking below these phenotypes. For *GRIN2A*, the strongest association was observed with developmental delays and disorders, followed by epilepsy. These findings highlight the strong specificity of epilepsy for *LGI1, ADAM23, GABRG2*, and *ATP1A3*, while genes such as *KRIT1, TSC1, TSC2, DEPDC5*, and *GRIN2A* exhibited additional associations reflective of their broader phenotypic impact.

## Discussion

Large population biobanks predominantly recruit middle-aged to older, well-educated individuals, leading to underrepresentation of individuals with developmental disabilities or severe early-onset conditions^13^. Consequently, epilepsy cases captured in these cohorts differ markedly from those in specialized genetic studies, which typically emphasize early-onset, severe, and syndromic forms of epilepsy. By analyzing a public dataset of rare coding variant associations in over 750,000 individuals derived from three independent biobanks, including 20,026 with epilepsy, we identified both established and novel epilepsy-associated genes.

These findings suggest that canonical epilepsy genes also contribute to susceptibility in milder or later-onset forms of the disease.

The results from our meta-analysis, compared to the largest exome study in epilepsy to date, the Epi25 exome sequencing study, reveal both overlapping and distinct genetic associations with epilepsy, highlighting methodological, phenotypic, and cohort differences^2^. Both studies reinforced the significance of *DEPDC5* and *GABRG2*. Differences likely arose from the broader phenotype in our study, which grouped diverse epilepsy presentations under a single phecode, and the subtype-specific analysis in the Epi25 study, which emphasized severe forms like developmental and epileptic encephalopathies. Our findings also emphasized the mTOR pathway (e.g., *DEPDC5, TSC1, TSC2*), while the Epi25 study highlighted additional pathways, such as ion channels and GABAA receptor complexes – many well-established severe pediatric epilepsy genes included^2^.

Notably, we identified a novel association with *ADAM23*, a gene previously linked to epilepsy in dogs^11^. *ADAM23* encodes a synaptic adhesion molecule that forms a complex with *LGI1*, the gene showing the strongest association in our study and a well-established epilepsy gene^12^. *LGI1* is implicated in a broad phenotypic spectrum, ranging from autosomal dominant lateral temporal lobe epilepsy to other forms of focal epilepsies. Its direct interaction partner, *ADAM23*, may similarly exhibit a wide—albeit subtler—range of epilepsy presentations. A homozygous knockout of ADAM23 leads to spontaneous seizures, while heterozygous mice exhibit a reduced seizure threshold. While the pathogenic role of *ADAM23* in human epilepsy remains to be confirmed, its identification in population-scale data raises the possibility of involvement in milder or later-onset forms of epilepsy, potentially modulated by additional genetic or environmental factors.

Phenome-wide association analyses revealed the pleiotropic effects of identified epilepsy genes. While *LGI1, ADAM23, GABRG2*, and *ATP1A3* were predominantly associated with epilepsy, genes such as *KRIT1, TSC1*, and *TSC2* demonstrated multisystemic involvement. These findings are consistent with prior evidence linking *KRIT1* to vascular phenotypes, including cerebral cavernous malformations, and associating *TSC1* and *TSC2* with congenital anomalies and arrhythmias characteristic of tuberous sclerosis complex^14, 15^. Similarly, *DEPDC5* was linked not only to neurological traits but also to unexpected phenotypes such as herpes zoster and uterine neoplasms, reflecting its broader biological roles. The association of *GRIN2A* with developmental delays further reinforces its contribution to neurodevelopmental disorders.

Our study has notable limitations. Phenotypes derived from billing codes lack the precision of specialist-defined cohorts, potentially leading to misclassification of epilepsy subtypes. The underrepresentation of individuals with developmental disabilities and refractory epilepsy likely biases findings toward milder cases. Furthermore, validating novel associations such as *ADAM23* will require functional studies and targeted recruitment. The focus on rare coding variants excludes noncoding and structural variants that may also influence epilepsy susceptibility. Despite these limitations, our reanalysis of population-scale data identified *ADAM23* as a candidate epilepsy gene not previously associated with the disorder in humans. Integrating genetic data from large-scale public resources with deep phenotyping, functional validation, and longitudinal follow-up will be essential for refining genotype–phenotype correlations and advancing personalized treatment strategies. Together, findings from population-based and research-ascertained cohorts are broadening our understanding of epilepsy genetics, moving beyond traditional syndromic boundaries.

## Data Availability

All data produced in the present work are contained in the manuscript

## Acknowledgement

Christian Bosselmann is supported by the MINT-Clinician Scientist program of the Medical Faculty Tübingen, funded by the Deutsche Forschungsgemeinschaft (DFG, German Research Foundation) – 493665037. Eduardo Pérez-Palma is supported by the Chilean National Agency for Investigation and Development (ANID), FONDECYT grant 1221464.

## REFERENCES

1. Globavl, regional, and national burden of neurological disorders, 1990-2016: a systematic analysis for the Global Burden of Disease Study 2016 Lancet Neurol. 2019 May;18:459–480.

2. Exome sequencing of 20,979 individuals with epilepsy reveals shared and distinct ultra-rare genetic risk across disorder subtypes Nat Neurosci. 2024 Oct;27:1864–1879.

3. Stefanski A, Calle-López Y, Leu C, Pérez-Palma E, Pestana-Knight E, Lal D. Clinical sequencing yield in epilepsy, autism spectrum disorder, and intellectual disability: A systematic review and meta-analysis Epilepsia. 2021 2021/01/01;62:143–151.

4. Boßelmann CM, Leu C, Brünger T, Hoffmann L, Baldassari S, Chipaux M, et al. Analysis of 1386 epileptogenic brain lesions reveals association with DYRK1A and EGFR Nat Commun. 2024 Nov 30;15:10429.

5. Heyne HO, Singh T, Stamberger H, Abou Jamra R, Caglayan H, Craiu D, et al. De novo variants in neurodevelopmental disorders with epilepsy Nat Genet. 2018 Jul;50:1048–1053.

6. GWAS meta-analysis of over 29,000 people with epilepsy identifies 26 risk loci and subtype-specific genetic architecture Nat Genet. 2023 Sep;55:1471–1482.

7. Wadhwa L. Landscape of Pediatric Biobanking: Challenges and Current Efforts Biopreserv Biobank. 2021 Apr;19:119–123.

8. Gramm M, Leu C, Pérez-Palma E, Ferguson L, Jehi L, Daly MJ, et al. Polygenic risk heterogeneity among focal epilepsies Epilepsia. 2020 Nov;61:e179–e185.

9. Jurgens SJ, Wang X, Choi SH, Weng LC, Koyama S, Pirruccello JP, et al. Rare coding variant analysis for human diseases across biobanks and ancestries Nat Genet. 2024 Sep;56:1811–1820.

10. Oliver KL, Scheffer IE, Bennett MF, Grinton BE, Bahlo M, Berkovic SF. Genes4Epilepsy: An epilepsy gene resource Epilepsia. 2023 May;64:1368–1375.

11. Koskinen LLE, Seppälä EH, Weissl J, Jokinen TS, Viitmaa R, Hänninen RL, et al. ADAM23 is a common risk gene for canine idiopathic epilepsy BMC Genetics. 2017 2017/01/31;18:8.

12. Yamagata A, Miyazaki Y, Yokoi N, Shigematsu H, Sato Y, Goto-Ito S, et al. Structural basis of epilepsy-related ligand–receptor complex LGI1–ADAM22 Nature Communications. 2018 2018/04/18;9:1546.

13. Fry A, Littlejohns TJ, Sudlow C, Doherty N, Adamska L, Sprosen T, et al. Comparison of Sociodemographic and Health-Related Characteristics of UK Biobank Participants With Those of the General Population American Journal of Epidemiology. 2017;186:1026–1034.

14. Denier C, Labauge P, Brunereau L, Cavé-Riant F, Marchelli F, Arnoult M, et al. Clinical features of cerebral cavernous malformations patients with KRIT1 mutations Annals of Neurology. 2004 2004/02/01;55:213–220.

15. Moloney PB, Cavalleri GL, Delanty N. Epilepsy in the mTORopathies: opportunities for precision medicine Brain Communications. 2021;3.

